# Novel approach to diagnosis of His bundle capture using individualized left ventricular lateral wall activation time as reference

**DOI:** 10.1101/2021.05.28.21257962

**Authors:** Marek Jastrzębski, Paweł Moskal, Piotr Kukla, Agnieszka Bednarek, Grzegorz Kiełbasa, Marek Rajzer, Karol Curila, Pugazhendhi Vijayaraman

**Affiliations:** First Department of Cardiology, Interventional Electrocardiology and Hypertension, Jagiellonian University Medical College, Kraków, Poland; Department of Cardiology, H. Klimontowicz Specialistic Hospital, Gorlice, Poland; Department of Cardiology, Third Faculty of Medicine, Charles University and University Hospital Kralovske Vinohrady, Prague, Czech Republic; Geisinger Heart Institute, Geisinger Commonwealth School of Medicine, Wilkes-Barre, PA, USA

**Keywords:** His bundle pacing, loss of capture, non-selective pacing, electrocardiogram

## Abstract

**Aims:** During non-selective His bundle (HB) pacing, it is clinically important to confirm His bundle capture vs. right ventricular septal (RVS) capture. The present study aimed to validate the hypothesis that during HB capture left ventricular lateral wall activation time, approximated by the V_6_ R-wave peak time (V_6_RWPT), will not be longer than the corresponding activation time during native conduction.

**Methods:** Consecutive patients with permanent HB pacing were recruited; cases with abnormal His-ventricle interval or left bundle branch block were excluded. Two corresponding intervals were compared: stimulus-V_6_RWPT and native HBpotential-V_6_RWPT. Difference between these two intervals (delta V_6_RWPT), diagnostic of lack of HB capture, was identified using receiver operating characteristic (ROC) curve analysis.

**Results:** A total of 723 ECGs (219 with native rhythm, 172 with selective HB, 215 with non-selective HB, and 117 with RVS capture) were obtained from 219 patients. The native HB-V_6_RWPT, non-selective-, and selective-HB paced V_6_RWPT were nearly equal, while RVS V_6_RWPT was 32.0 (±9.5) ms longer. The ROC curve analysis indicated delta V_6_RWPT > 12 ms as diagnostic of lack of HB capture (specificity of 99.1% and sensitivity of 100%). A blinded observer correctly diagnosed 96.7% (321/332) of ECGs using this criterion.

**Conclusion:** We validated a novel criterion for HB capture that is based on the physiological left ventricular activation time as an individualized reference. HB capture can be diagnosed when paced V_6_RWPT does not exceed the value obtained during native conduction by more than 12 ms, while longer paced V_6_RWPT indicates RVS capture.

## Introduction

His bundle (HB) pacing is a new method of permanent ventricular pacing, still far from complete electrocardiographic and electrophysiological characterization. It is particularly important to develop accurate criteria for the electrocardiographic diagnosis of HB capture / loss of HB capture during follow-up. During HB pacing, simultaneous activation of the adjacent right ventricular septal (RVS) myocardium often occurs, resulting in non-selective (ns-)HB capture. In such a situation, loss of HB capture might be masked by the still maintained RVS capture.^1^ This makes ECG-based diagnosis of HB capture challenging. In most patients with non-selective HB capture, output dependent changes in QRS morphology is used to confirm HB capture. However, in approximately 8-10% of patients, both HB and RVS capture thresholds are identical where accurate diagnosis of HB capture can be difficult. To address this problem, an ECG algorithm was developed that is based on the observation that the V_6_ R-wave peak time (V_6_RWPT), a parameter that corresponds with the activation time of the left ventricular (LV) lateral wall, is longer during RVS capture than during ns-HB capture.^2^ However, LV activation times during both physiological and non-physiological ventricular depolarization show typical bell curve distribution, leading to some overlap of values observed during RVS capture and ns-HB capture.^2^ Therefore, any fixed cut-off QRS-based duration criterion for diagnosis of HB capture is significantly limited by a trade-off between the sensitivity and specificity.

The present study was prompted by the hypothesis that conduction system capture should be characterized by equal left ventricular (LV) lateral wall activation times during pacing and during native conduction. Therefore, using native LV lateral wall activation time (that is V_6_RWPT) as reference instead of fixed cut-offs allows individualization of duration criteria. Such an approach was already proposed and validated for the diagnosis of left bundle branch capture during deep septal pacing.^3^ In the current study, we aimed to validate the above hypothesis in the setting of HB pacing. Specifically, we hypothesized that during HB capture, the pacing stimulus to the V_6_ R-wave peak (paced V_6_RWPT) should be nearly equal to the HB potential to the V_6_ R-wave peak (HB-V_6_RWPT), while lack of HB capture, that is, only RV septal myocardial capture, should be characterized by longer paced V_6_RWPT than HB-V_6_RWPT. This principle is illustrated in Figures 1 and 2.

**Figure 1.**
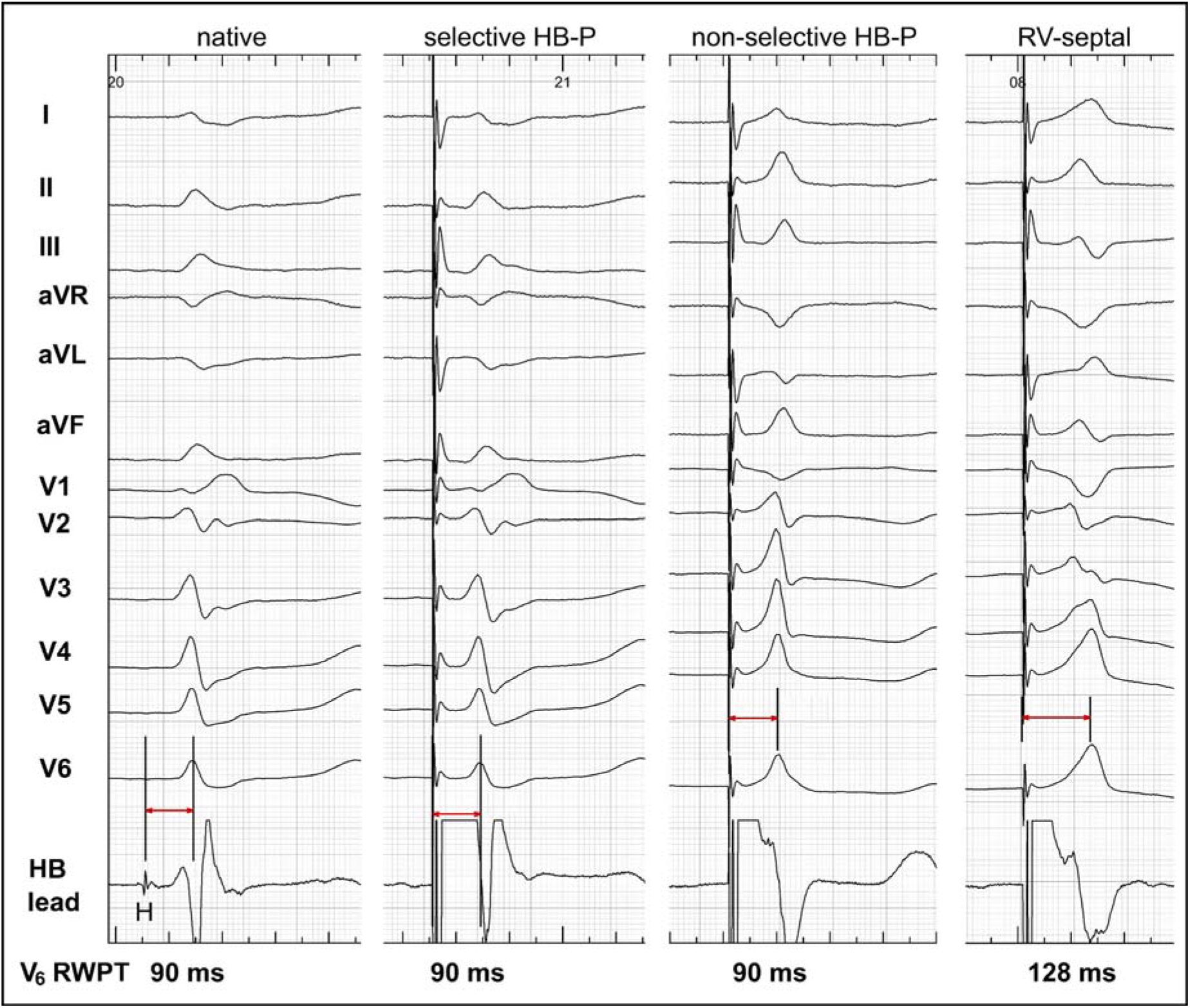
Activation time of the lateral wall of the left ventricle, as indicated by the time to the R-wave peak in lead V6 (V_6_RWPT), remains constant during native conduction and His bundle pacing (HB-P). Loss of HB capture, that is right ventricular (RV) septal-only capture, results in prolongation of paced V_6_RWPT by 38 ms (from 90 ms to 128 ms). In contrast to the lateral wall activation time, the global ventricular activation (i.e. activation time till the QRS offset) cannot serve in this case as reference since RV septal QRS is narrower than HB potential to QRS offset interval during native conduction (178 ms vs. 206 ms, respectively).

**Figure 2.**
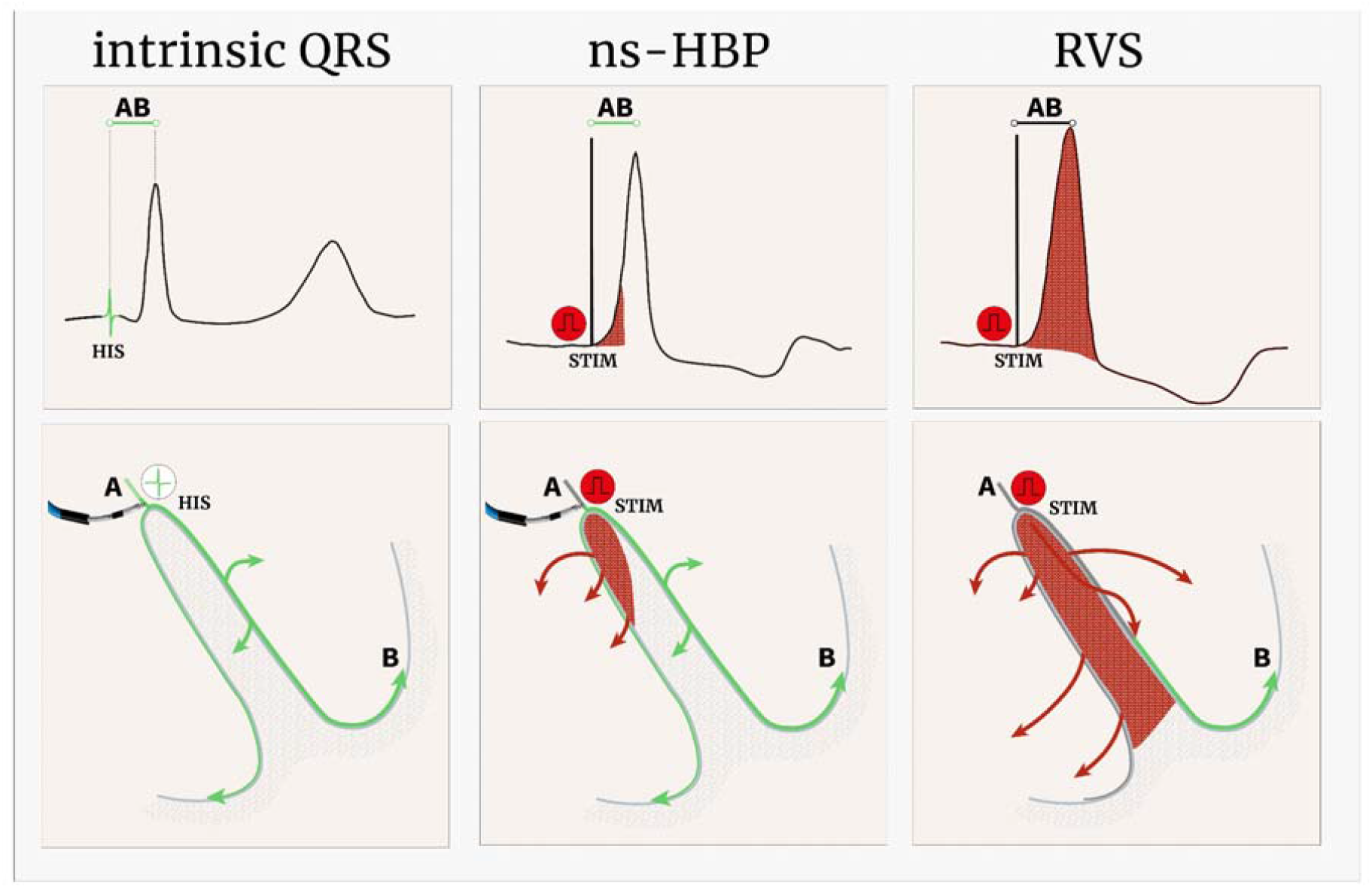
Schematic illustration of the equal left ventricular activation time criterion for the diagnosis of His bundle capture. During intrinsic activation, the time from the His bundle potential (HIS) to the activation of the lateral wall of the left ventricle equals the time from the pacing stimulus (STIM) to the activation of the lateral wall of the left ventricle during non-selective HB pacing (ns-HBP). During only right ventricular septal (RVS) capture, the corresponding interval is longer by the time it takes for the depolarization wavefront to cross the interventricular septum and engage the conduction system of the left ventricle. Comparison of the paced AB interval with the intrinsic AB interval enables accurate differentiation between ns-HB capture and RVS capture.

## Methods

The data that support the findings of this study are available from the corresponding author upon reasonable request. The study adhered to the Helsinki Declaration as revised in 2013 and was approved by the Institutional Bioethical Committee.

### Population

We screened all consecutive patients who underwent conduction system pacing device implantation for bradycardia and/or heart failure indications in our center in the years 2014– 2021. The following inclusion criteria were applied for the current study:

1. His bundle potential was recorded at the final implantation site of the pacing lead.
2. The His-ventricle (HV) interval was within the normal range of 35–55 ms.
3. The intrinsic supraventricular QRS was of non-left bundle branch block type (per Strauss definition).
4. HB capture was confirmed with the gold standard method, i.e. demonstration of QRS morphology change due to transition from ns-HB capture to either selective (s-)HB capture or RVS myocardial capture. QRS morphology transition was obtained either during capture threshold (output dependent transition) and/or programmed stimulation (refractoriness dependent transition).

### Definitions and measurements

In every studied patient, each available paced QRS type (s-HB, ns-HB and RVS), as well as QRS during the native supraventricular rhythm, was analyzed. The following QRS characteristics were obtained:

1. Paced V_6_RWPT (interval from pacing stimulus to R-wave peak in lead V6)
2. HV interval (interval from His bundle potential to QRS onset)
3. HB-V_6_RWPT (interval from His bundle potential to R-wave peak in lead V6)
4. Global QRS duration (interval from the earliest onset to the latest offset of the QRS in all 12 leads recorded simultaneously; for paced QRS, pacing stimulus was considered as QRS onset).

All implantation procedures were recorded on the digital electrophysiological system (LabsystemPRO, Boston Scientific, USA). His bundle potential was recorded by connecting the HB pacing lead to a separate channel in the electrophysiological system using alligator clips; the high-pass filter for this channel was set at 30 Hz and the low-pass filter was set at 100 Hz. To ensure high precision of measurements all 12 surface ECG leads and the endocardial channel were recorded simultaneously and digital calipers, fast sweep speed (200 mm/s), and appropriate signal augmentation were used. At least three QRS complexes/intervals were measured and their values averaged.

### Statistical analysis

Continuous variables are presented as means and standard deviations. Distribution of the HB-V_6_RWPT, ns-HB V_6_RWPT, s-HB V_6_RWPT, RV-septal V_6_RWPT, and QRS duration during all forms of ventricular activation was estimated by the kernel method. Categorical variables are presented as percentages. The performance of binary decision rules is described using sensitivity (SN) and specificity (SP). The activation times were compared using dependent Student’s t-test. Pearson’s correlation was used to determine the relation between the native activation time (HB-V_6_RWPT) and the corresponding paced activation time (ns-HB V_6_RWPT). Receiver operating characteristic (ROC) curve analysis was used to determine optimal cut-off for QRS duration, V_6_RWPT, and the difference between native and paced activation (delta V_6_RWPT, delta QRS). To compare the area under the curve (AUC) of two paired ROC curves, the DeLong approach was used. Statistical analyses were performed using R software (The R Foundation). P-values < 0.05 were considered statistically significant.

### Validation of the practical application of the ‘equal activation times’ criterion

All ECGs with ns-HB capture and RVS capture were saved (jpg graphic files) with standard 12-lead ECG with paper speed of 25 mm/s and deidentified using labels with random numbers. A blinded observer (a general cardiologist not involved with conduction system pacing) was asked to make a diagnosis regarding the type of capture, namely ns-HB or RVS, just on the basis of the provided HB-V_6_RWPT value for a particular patient (that was obtained at the end of the implantation procedure) using the ROC-based ‘equal activation times’ criterion. He analyzed ECG traditionally and without help of digital calipers albeit could enlarge the tracings on computer monitor to facilitate interval measurements.

## Results

### Population

A total of 1042 patients who had undergone conduction system pacing device implantation were screened, and 219 patients were identified who fulfilled the inclusion criteria. These 219 patients provided 723 ECGs for analysis, including 215 ECGs with ns-HB capture, 219 ECGs with native rhythm, 172 ECGs with s-HB capture and 117 ECGs with RVS capture. Basic clinical characteristics of the studied group are presented in Table 1.

**Table 1.** Basic characteristics of the studied group (n = 219)

|  |  |
| --- | --- |
| Age [years] | 75.4 ± 10.7 |
| Male gender | 131 (59.8%) |
| Pacing indication [n]: |  |
| • Sick sinus syndrome | 55 (25.1%) |
| • Atrioventricular block | 59 (26.9%) |
| • Atrial fibrillation with bradycardia | 187 (33.3%) |
| • Heart failure | 32 (14.6%) |
| Comorbidities [n]: |  |
| • Diabetes mellitus | 77 (35.2%) |
| • Coronary heart disease | 71 (32.4%) |
| • Heart failure | 89 (40.6%) |
| • Hypertension | 187 (85.4%) |
| • Severe valvular disease | 16 (7.3%) |
| Left ventricular ejection fraction [%] | 51.4 ± 12.7 |
| Left ventricular end-diastolic dimension [mm] | 50.6 ± 7.8 |
| His- ventricle interval [ms] | 43.5 ± 5.6 |
| Native QRS duration [ms] | 113.5 ± 22.0 |
| Native QRS type: |  |
| • Narrow | 75 (34.2%) |
| • LAFB | 8 (3.6%) |
| • Incomplete RBBB | 4 (1.8%) |
| • RBBB | 73 (33.3%) |
| • RBBB + LAFB/LPFB | 16 (7.3%) |
| • NIVCD | 43 (19.6%) |
| Acute capture threshold [V] | 1.28 ± 0.7 |
| Acute ventricular sensing [mV] | 3.7 ± 2.3 |

RBBB – right bundle branch block; LAFB – left anterior fascicular block; LPFB – left posterior fascicular block; NIVCD – non-specific intraventricular conduction disturbance.

The reasons for exclusion of the remaining 823 patients were as follows: left bundle branch pacing (n = 489), HB pacing not fulfilling the inclusion criteria (n = 85), right bundle branch pacing (n = 83), and conduction system pacing failures/miscellaneous reasons (n = 166).

### Lead V_6_ R-wave peak time criteria

The comparison of V_6_RWPT intervals between different types of ventricular depolarization is presented in Table 2 and Figure 3. Native activation time (HB-V_6_RWPT) and paced activation time during non-selective capture (ns-HB V_6_RWPT) were nearly equal (92.4 ±10.4 ms vs. 92.9 ±10.6 ms, p = 0.08), with a very strong correlation between them (r = 0.921, p < 0.001) (Figure 4). In contrast, the activation time during native conduction compared to RVS capture varied, with an average difference of 33.9 ms (93.0 ±10.0 ms vs. 126.9 ±13.8 ms, p < 0.001). Despite this difference, there was still a considerable overlap of activation time values between RVS and ns-HB capture, as illustrated in the density plots (Figure 2, upper panels). However, using native conduction as a reference, which enabled calculation of delta V_6_RWPT, resulted in nearly complete separation of ns-HB capture and RVS capture (Figure 3).

**Table 2.**
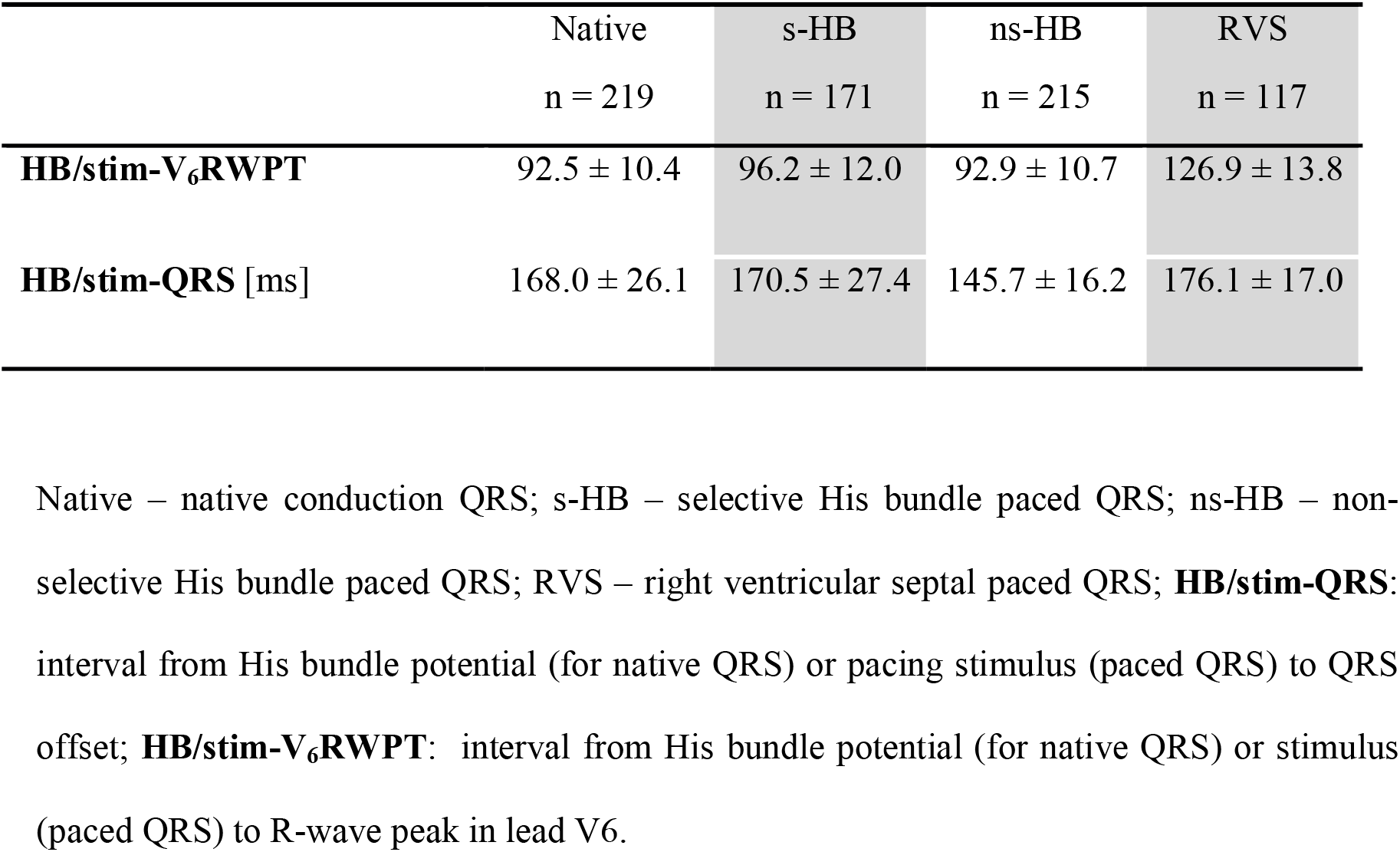
Comparison of duration parameters during pacing and native conduction

|  | Native<br>n = 219 | s-HB<br>n = 171 | ns-HB<br>n = 215 | RVS<br>n = 117 |
| --- | --- | --- | --- | --- |
| **HB/stim-V<sub>6</sub>RWPT** | 92.5 ± 10.4 | 96.2 ± 12.0 | 92.9 ± 10.7 | 126.9 ± 13.8 |
| **HB/stim-QRS [ms]** | 168.0 ± 26.1 | 170.5 ± 27.4 | 145.7 ± 16.2 | 176.1 ± 17.0 |

Native – native conduction QRS; s-HB – selective His bundle paced QRS; ns-HB – non-selective His bundle paced QRS; RVS – right ventricular septal paced QRS; **HB/stim-QRS**: interval from His bundle potential (for native QRS) or pacing stimulus (paced QRS) to QRS offset; **HB/stim-V<sub>6</sub>RWPT**: interval from His bundle potential (for native QRS) or stimulus (paced QRS) to R-wave peak in lead V6.

**Figure 3.**
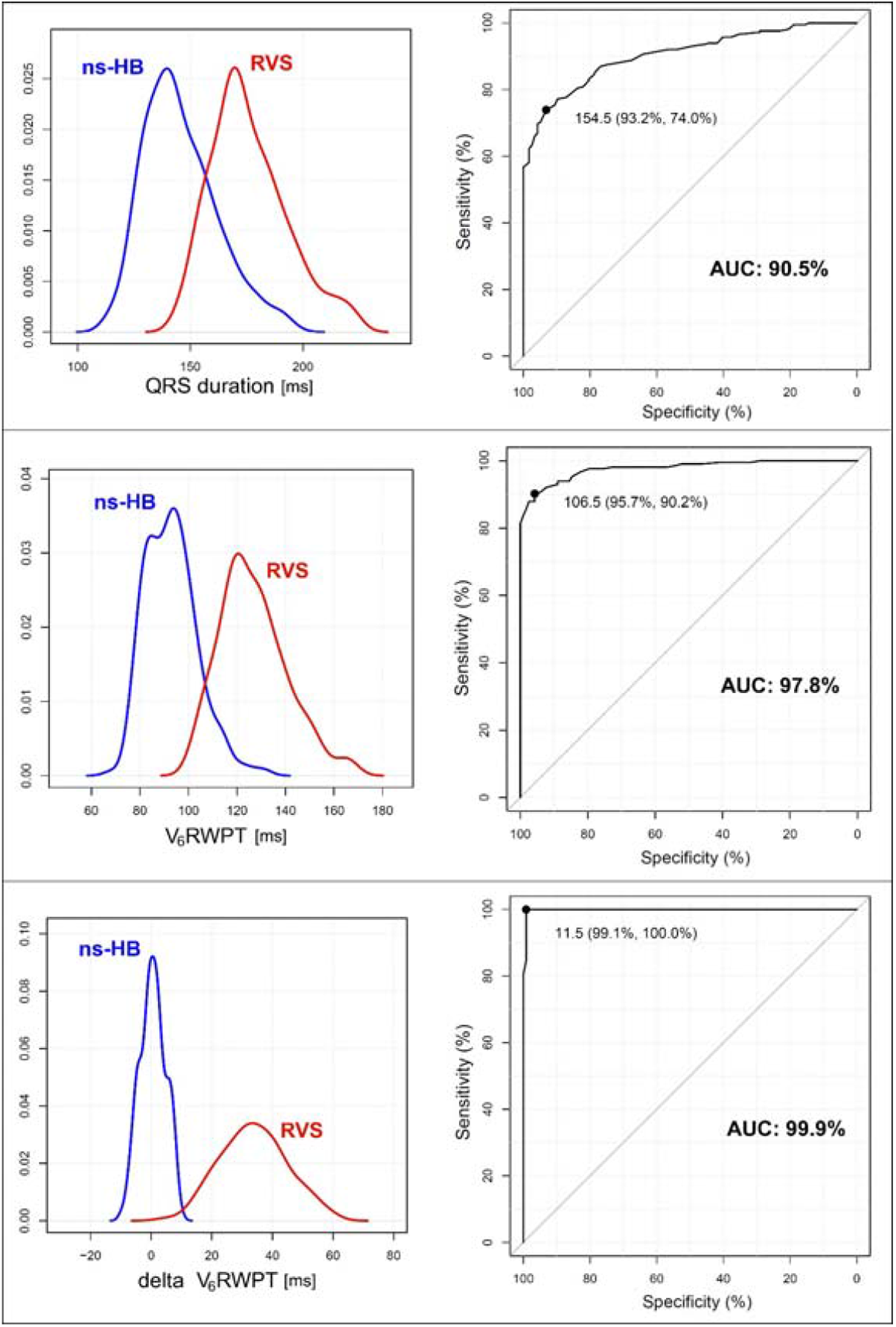
Density plots and corresponding receiver operating characteristics curves for QRS duration, R-wave peak time in lead V_6_ (V_6_RWPT) and delta V_6_RWPT during non-selective His bundle capture (ns-HB) and right ventricular septal (RVS) capture. There is progressively smaller overlap of values between ns-HB and RVS capture and growing diagnostic performance as expressed by the area under the curve (AUC) with each subsequent criterion (p < 00.1). Values in parentheses represent specificity and sensitivity, respectively.

**Figure 4.**
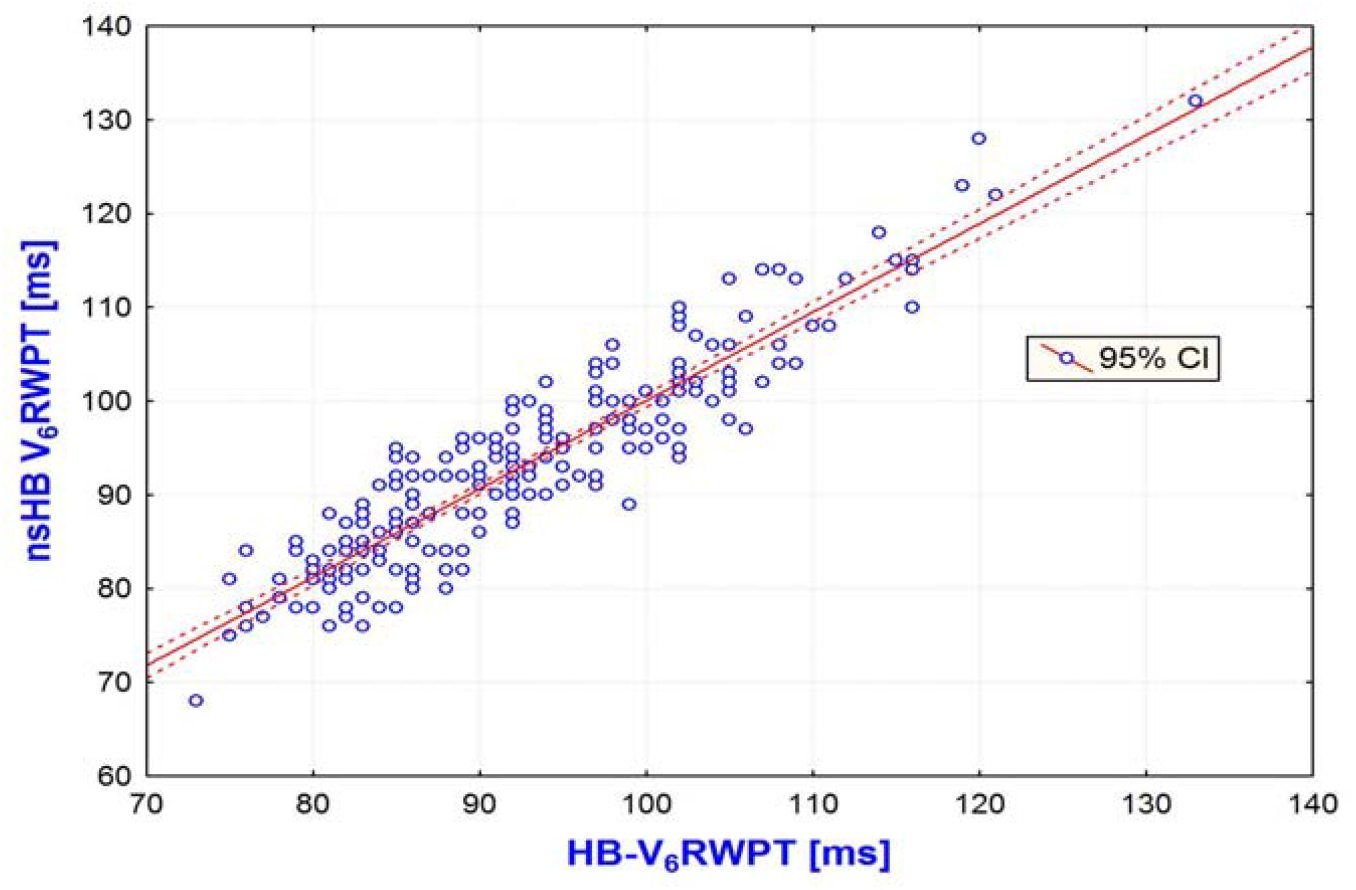
Correlation between native activation time (HB-V_6_RWPT) and paced activation time (ns-HB V_6_RWPT). CI – confidence intervals. R = 0.921, R^2^ = 0.849; p < 0.001

The results of ROC curve analysis, presented in Figure 3, pointed to a difference between HB-V_6_RWPT and paced V_6_RWPT (delta V_6_RWPT) of less than 12 ms as an optimal criterion for ns-HB capture diagnosis. This criterion was characterized by area under the ROC curve of 99.9%, and SN, SP and accuracy of the diagnosis of HB capture of 100%, 99.1% and 99.7%, respectively.

Diagnostically optimal fixed cut-off V_6_RWPT value was identified as 106.5 ms (Figure 3). The area under the ROC curve, SN, and SP of the fixed cut-off V_6_RWPT criterion were 97.8%, 90.2% and 95.7%, respectively.

Comparison of the area under the ROC of 99.9%, 97.8%, and 90.5% for delta V_6_RWPT, fixed cut-off V_6_RWPT, and QRS duration, respectively, revealed significant differences between each pair (p < 0.01).

### Global QRS duration criteria

Comparison of global QRS duration-based intervals between different types of ventricular depolarization is presented in Table 2 and Figure 3. Native global ventricular activation time (HB-QRS offset) and paced global ventricular activation time during selective HB capture were nearly equal, with an average difference of 2.5 ms (168.0 ±25.9 ms vs. 170.5 ±27.4 ms, p < 0.009), while the global ventricular activation during ns-HB capture and RV septal capture was faster by 22.2 ms (167.9 ±26.1 ms vs. 145.7 ±16.2 ms, p < 0.001) and slower by 7.3 ms (168.8 ±26.3 ms vs. 176.1 ±17.0 ms, p < 0.004), respectively. Density plots showed considerable overlap of global ventricular activation time values (QRS duration, Figure 2) between ns-HB and RVS capture. Diagnostically optimal fixed cut-off for QRS duration was identified by the ROC curve analysis as 154.5 ms. This criterion was characterized by an SN of 74.0% and an SP of 93.2% for the diagnosis of HB capture.

Using native conduction as reference (delta QRS) was not helpful, since it worsened the separation of ns-HB capture and RVS capture values (Supplemental Figure 1) and decreased the area under the ROC curve from 90.5% to 77.5%.

This results prompted post-hoc subgroup analysis of global QRS fixed cut-off and delta QRS criteria in patients with narrow QRS (Supplemental Figure 1). In patients with narrow QRS both these criteria performed nearly as well as V_6_RWPT fixed cut-off and delta V_6_RWPT. Diagnostically optimal fixed cut-off QRS duration in narrow QRS patients value was identified as 152.5 ms (Supplemental Figure 1). The area under the ROC curve, SN, and SP of the fixed cut-off V_6_RWPT criterion were 97.8%, 86.0% and 95.6%, respectively. Diagnostically optimal fixed delta QRS duration value in narrow QRS patients was identified as 7.5 ms (Supplemental Figure 1). The area under the ROC curve, SN, and SP of the delta QRS criterion were 98.6%, 95.2% and 93.2%, respectively.

### Validation of practical application of the individualized lead V6 R-wave peak time criterion

A total of 332 ECGs (117 with RVS and 215 with ns-HB capture) were analyzed by a blinded observer who was provided with HB-V_6_RWPT values for each patient. A correct diagnosis of RV or ns-HB capture was made by this observer in 115/117 RVS ECGs and in 206/215 ns-HB capture ECGs, resulting in a sensitivity of 95.8% and a specificity of 98.3% for diagnosis of HB capture.

## Discussion

The research hypothesis that HB capture reproduces native, physiological activation times in the left ventricle was positively validated. The key finding of our study, based on this principle, was that the left ventricle lateral wall activation time during native conduction (HB-V_6_RWPT) can serve as a reference for confirmation of HB capture in patients with normal HV interval. The individualized V_6_RWPT criterion (delta V_6_RWPT) resulted in diagnostic accuracy of 99.7% that surpassed the fixed V_6_RWPT cut-off criterion and QRS duration-based criteria.

### Physiological perspective

The V_6_RWPT corresponds to the intrinsicoid deflection time in lead V6, reflecting the time it takes for the depolarization wavefront to reach the epicardial surface of the lateral wall of the LV.^4, 5^ During ns-HB pacing, the physiological activation of the LV remains largely unaffected (**Figure 2**), despite direct non-physiological septal activation that leads to pseudo-delta formation and QRS prolongation that characterizes ns-HB pacing. This is because the velocity of conduction in the His-Purkinje system is much higher than in the septal myocardium. With the possible exception of basal segments of the interventricular septum and the posterior wall,^6^ the rest of the LV, particularly the lateral wall, is depolarized purely via the His-Purkinje conduction system. Consequently, during ns-HB pacing, the time from the pacing stimulus (that corresponds to HB depolarization during native conduction) to the activation of the lateral wall of the LV is not influenced by the direct myocardial capture of the septum and should be nearly equal to the corresponding time during native supraventricular conduction (HB-V_6_RWPT = stimulus-V_6_RWPT). The results of the present study fully corroborate the above reasoning, adding to the previously published data that physiological LV activation times are the hallmarks of physiological pacing.^2, 3^ We believe that longer LV activation times could be mechanistically interpreted as the degree of deviation from physiology during pacing.

Using global QRS duration for diagnosis of HB capture is also possible,^7^ albeit, as shown in this study and in our previous research,^2^ it is inferior to V_6_RWPT because of bigger overlap of QRS duration values between ns-HB and RVS capture. Global QRS duration reflects right and left ventricular activation and not only LV lateral wall activation. Therefore, in contrast to V_6_RWPT, global QRS duration <u>is</u> influenced by direct myocardial capture during RVS and ns-HB pacing. This is especially true in patients with right bundle branch block (Figure 1). During right bundle branch block, RVS capture partially corrects delay in RV activation, resulting in QRS narrowing. Consequently, RVS paced QRS might be narrower than the HB-QRS offset interval during native conduction (Figure 1) - limiting the application of QRS based criteria to patients with narrow QRS (Supplementary Figure 1). Moreover, using global QRS duration as a criterion is also limited by the inaccuracies of measurement; QRS offset, in contrast to R-wave peak, is not so easy to determine precisely and reproducibly. We believe that these were the reasons for the smaller area under the ROC curve for the QRS-based criteria in comparison to the V_6_RWPT criteria.

### Clinical implications

Diagnosis of HB capture during the implantation procedure is easy in the vast majority of cases, since the HB capture threshold is usually different from the RVS capture threshold. This makes it possible to observe diagnostic change of QRS morphology during decrease in pacing output. However, in a small percentage of patients, the intraprocedural capture thresholds are equal and then, for the diagnosis of HB capture, different maneuvers have to be used, like programmed stimulation or burst pacing, that exploit differences in refractoriness between HB and septal myocardium.^8, 9^ The currently developed method offers a faster and simple alternative for the determination of HB capture in cases where the threshold test fails as a diagnostic test. Potentially, it can be even used as a primary diagnostic approach since the His bundle potential is almost always recorded at the implantation site and precise measurement of native HB-V_6_RWPT and paced V_6_RWPT is straightforward.

The currently developed method/criterion might be even more useful during follow-up of patients with HB pacing devices. In the setting of a non-device clinic, when the device programmer is not available, the diagnosis of proper pacing, that is HB capture / loss of HB capture, has to be based on ECG interpretation. Both pacing stimulus and R-wave peak are very clear markers (in contrast to QRS onset and offset) and allow precise determination of paced V_6_RWPT even on the basis of a standard 12-lead ECG recorded with a 25-mm/s paper speed. This was corroborated by the validation part of the current research, where a general cardiologist was able to correctly diagnose HB capture/non-capture with an overall accuracy of 96.7%.

There is one important caveat: using this criterion during follow-up requires knowledge of the reference value, namely the HB-V_6_RWPT obtained during the implantation procedure. Perhaps the HB-V_6_RWPT (or paced V_6_RWPT) value should be routinely included in the procedure report along with the capture thresholds, sensitivity values, and other relevant data. Importantly, monitoring the stability of V_6_RWPT during follow-up might reveal not only loss of HB capture but also the development of an intrahisian or intra-left ventricular conduction disturbance. Such a new conduction disturbance might be masked by the still present ns-HB capture, yet it may limit or abolish the physiological depolarization of the LV and the very purpose of HB pacing.

## Limitations

Our study group was limited to patients with normal HV interval. In patients with asystole / escape ventricular rhythm, native LV lateral wall activation time cannot be used as reference. Moreover, HB pacing can correct prolonged conduction in the HB, leading to the shortening of HV interval and/or correct left bundle branch block, which in turn results in the shortening of V_6_RWPT. Therefore, in patients with prolonged HV interval and/or LBBB, the native LV conduction times and native QRS cannot serve as diagnostic reference. In the current study, such patients (escape ventricular rhythm, HV > 55 ms, or LBBB), who constituted 22.4% of the whole population of patients with HB pacing devices, were excluded. However, this is a practical limitation rather than a methodological limitation of the current research; for such patients, the proposed fixed cut-off V_6_RWPT criterion or other methods should be used. Nevertheless, even in such cases, measurement and comparison of the native HB-V_6_RWPT and paced V_6_RWPT might still provide important insights. When, for a patient with prolonged HV interval and/or LBBB, the HB-V_6_RWPT equals paced V_6_RWPT, it clearly indicates that the underlying conduction problem was not corrected and a more distal pacing site in the conduction system should probably be explored. But if the paced V_6_RWPT is shorter than the baseline HB-V_6_RWPT, this suggests that the intrahisian and/or intra-left ventricular conduction disturbance was partially or fully corrected.

## Conclusions

A novel criterion for the diagnosis of HB capture has been formulated, which makes it possible to diagnose HB capture during presumed non-selective pacing when paced V_6_RWPT does not exceed the reference activation time obtained during native conduction by more than 12 ms; longer paced V_6_RWPT points to RVS capture. This criterion allowed accurate diagnosis in approximately 99% of cases and might be useful both at the time of implantation and during electrocardiographic-only follow-up.

## Data Availability

Data avilable upon reasonable request

