## Supplemental Figure 1 for "Novel approach to diagnosis of His bundle capture using individualized left ventricular lateral wall activation time as reference"

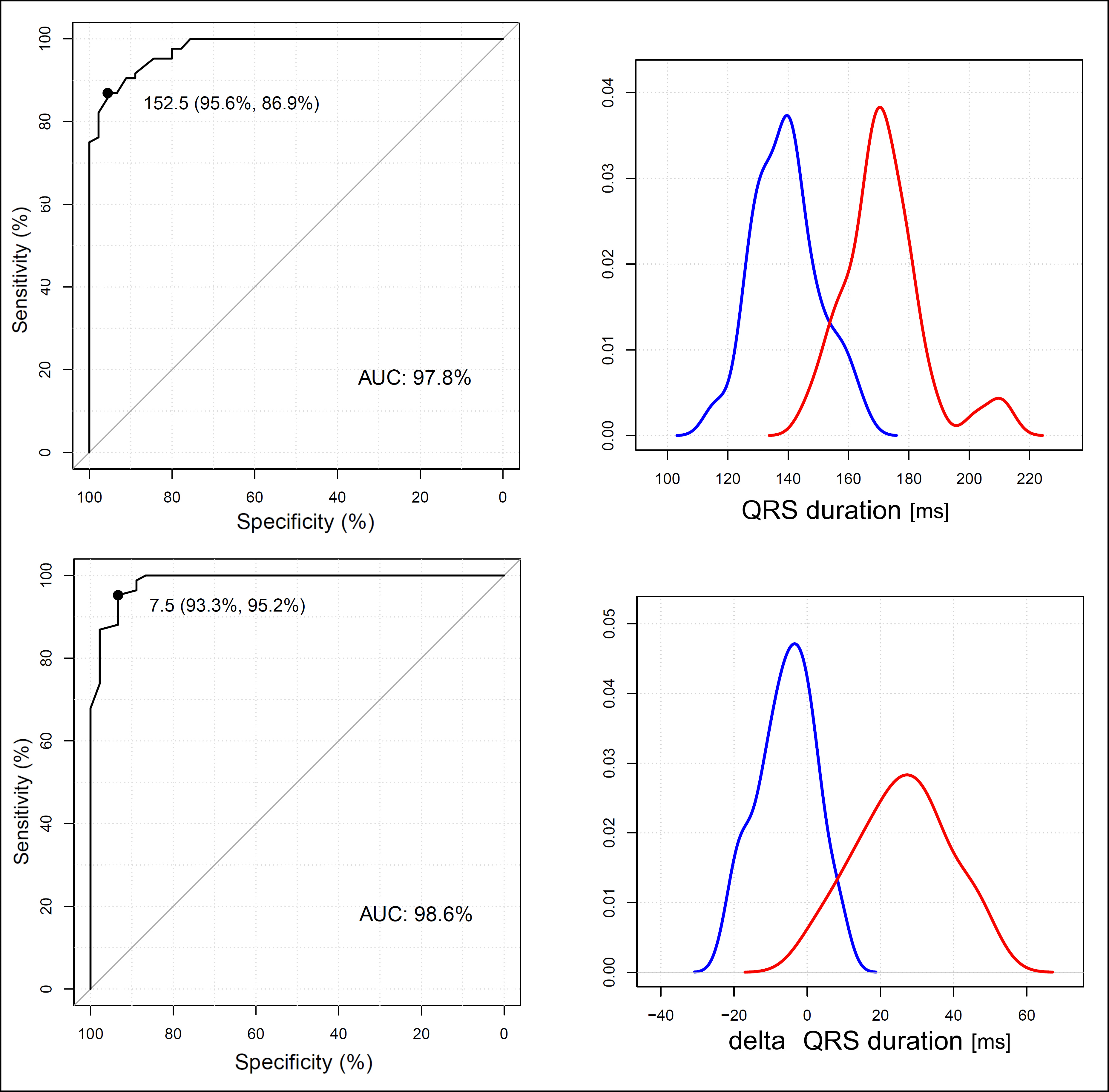

**Supplementary Figure 1.** Density plots and corresponding receiver operating characteristics curves for QRS duration and delta QRS duration in patients with narrow intrinsic QRS complexes. AUC - area under the curve. Values in parentheses represent specificity and sensitivity, respectively.
